# Assessment of Deep Learning-Based Triage Application for Acute Ischemic Stroke on Brain MRI in the Emergency Room

**DOI:** 10.1101/2023.10.05.23296577

**Authors:** Jimin Kim, Se Won Oh, Jee Young Kim, Heiko Meyer, Stefan Huwer, Gengyan Zhao, Dongyeob Han

**Affiliations:** Department of Radiology, Eunpyeong St. Mary’s Hospital, The Catholic University of Korea College of Medicine, Seoul, 03312, Korea; Siemens Healthcare GmbH, Erlangen, Henkestraße 127, 91052, Germany; Siemens Medical Solutions USA, Inc., Princeton, NJ 08540, United States of America; Siemens Healthineers Ltd., Seoul, 03737, Korea

**Keywords:** Acute ischemic stroke, Artificial intelligence, Deep learning, Triage research application, MRI, DWI, NeuroTriage

## Abstract

**Backgrounds:** To improve the outcomes of acute ischemic stroke (AIS) patients, a well-organized triage system is essential in the emergency room (ER). This study aimed to assess the detection performance of a deep learning triage research application for AIS that was newly developed based on brain MRI in the ER.

**Methods:** This retrospective study consecutively enrolled 831 brain MRIs including mandatory diffusion-weighted image (DWI) performed in the ER of our institution from April to October 2021. MRIs were analyzed with this triage research application as the index test and results were compared with the gold standard of three neuroradiologists. We evaluated sensitivity, specificity, area under the receiver operating characteristic curve (AUROC), area under the precision-recall curve (AUPRC), and maximum F1 score for this application. We also compared changes in its detection performance with and without the addition of optional sequences, which were T1- and T2-weighted images.

**Results:** 831 individuals (mean age, 64 years ± 16; 399 men, 432 women) were enrolled and 201 were positive for AIS. The application showed detection performance as follows; sensitivity, 90%; specificity, 89%; AUROC, 0.95 (95% confidence interval, 0.93-0.96); AUPRC, 0.91 (95% confidence interval, 0.86-0.94); and maximum F1 score, 0.86. The addition of optional sequences led to inferior performance compared to the mandatory sequence alone for detecting AIS.

**Conclusions:** The triage research application accurately detected AIS in a real-world ER with no additional benefits to its detection performance with optional sequences. Therefore, the triage research application can potentially help clinicians detect AIS in the ER sufficiently/adequately with only mandatory DWI.

## Introduction

Stroke is the second leading cause of mortality and disability worldwide and ischemic stroke accounts for approximately 87% of all strokes ^1-3^. Comprehensive early diagnosis and reperfusion treatment for acute ischemic stroke (AIS) is crucial to improving the long-term outcomes of AIS patients ^2,4^. Therefore, a well-organized triage system that can hasten early diagnosis is essential for a more favorable prognosis of AIS patients in the emergency room ^5,6^.

The artificial intelligence-based triage research application (NeuroTriage Research Application, version 1.2.5, Siemens Healthineers, Erlangen, Germany) to be discussed in this study was developed for the automated detection of critical findings of stroke including AIS. It utilized a deep learning 3D neural network system based on multi-parametric brain MRI of diffusion-weighted images (DWI) with an apparent diffusion coefficient (ADC) map for detecting AIS. A previous study that performed internal and external validation tests for this triage research application showed promising results ^7^. However, the prior validation study was not based on real-world scenarios that can occur in clinical settings.

The objective of our study was to evaluate the performance of the triage research application for detecting AIS in the emergency room. Additionally, we compared the detection performance of this application when the mandatory sequence was used alone and when optional sequences were added.

## Methods

This retrospective study was conducted according to the guidelines of the Declaration of Helsinki and approved by the Institutional Review Board of Eunpyeong St. Mary’s Hospital, The Catholic University of Korea College of Medicine (Protocol number PC22RESI0057, approval date March 30, 2022). The Institutional Review Board waived the requirement for informed consent.

### Study Sample

We consecutively enrolled 1224 patients visiting our institution’s emergency department who underwent a brain MRI examination under suspicion of AIS from April to October 2021. Every individual underwent one MRI examination. These examinations were reviewed by three board-certified neuroradiologists (J. K., S.-W.O., and J.-Y.K. with 8, 16, and 21 years of experience in brain MRI interpretation, respectively) to ascertain their eligibility for the final study population. 391 examinations were excluded due to the absence of the mandatory DWI sequence required for the triage research application analyses, and 2 other examinations were also excluded due to poor image quality caused by metal artifacts and acute ischemia in the cervical cord. Finally, 831 examinations were included for the triage research application image analyses. The data of these brain MRI examinations were uploaded to a server other than the application server and retrospectively analyzed. The same three neuroradiologists (J. K., S.-W.O., and J.-Y.K.) established a gold standard for acute AIS. The reviewers were blinded to each patient’s clinical information, previous readings, or each other’s reports at the initial reading. If the reviewers disagreed, the final decision about a brain MRI was then made in consensus. The sample size of this study was determined statistically feasible based on data from a previous study with a statistical significance of 0.05 and a statistical power of 0.80 ^7^. Further details on the estimation of sample size are provided in the <u>Data Supplement</u>.

### Diagnosis of AIS and lacunar AIS on MRI

We defined an AIS as a cerebral region with restricted diffusion due to cytotoxic edema in the brain parenchyma that is indicated by hyperintense signals on DWI with a corresponding low value on the ADC map ^8^. We also defined lacunar AIS noncortical infarcts less than 15 mm in diameter in the distal distribution of deep penetrating vessels, such as the lenticulostriate and thalamoperforating artery territories ^9,10^. Figure S1 in the <u>Data Supplement</u> presents examples of AIS and potential artifacts mimicking AIS.

### Network architecture and development of the triage research application

This application was developed utilizing a deep learning 3D convolutional neural network architecture for AIS, acute hemorrhagic stroke, and substantial mass effect. Using brain MRI inputs, the model calculates the percent probabilities for each incident’s critical findings. The application consists of three components; localization feature extraction modules, which are deep image-to-image networks with convolution and pooling layers; orientation-specific feature connection layers, which make the network robust to missing contrasts; and a global classifier that generates a probability score based on the presence of critical findings. To detect and determine the AIS network architecture, an axial DWI with the ADC map is mandatory, and axial T1- and T2-weighted images are optional. The network architecture is briefly illustrated in Figure S2 in the <u>Data Supplement</u>.

During its development, this triage research application utilized a total of 12,143 brain MRI studies for training (74%), tuning (9%), internal validation (8%), and external validation (8%). The flowchart of the development and validation for the triage research application is presented in Figure S3 in the <u>Data Supplement</u>. Additional heat maps were generated for each contrast to indicate the significance of the input pixel, and the localization feature extraction module generated these maps with pixel-wise significance in the input space ^7^.

According to Nael et al., a critical finding can be interpreted as positive if the triage research application calculates its risk probability as greater than 50% ^7^. Previous results for detecting AIS with this application are summarized in Table S1 in the <u>Data Supplement.</u>

### MRI protocol

All patients were scanned on a 3T MRI scanner (MAGNETOM Vida, Siemens Healthineers, Erlangen, Germany) with a 64-channel head & neck coil. In every examination, a standard infarct MRI protocol that included the axial DWI was carried out. The DWI was acquired with the following parameters using readout-segmented echo-planar diffusion: repetition time, 3720 ms; echo time, 61 ms; generalized auto-calibrating partial parallel acquisition factor, 2; number of excitation, 1; matrix size, 180 ⨯ 180; flip angle, 180 °; field of view, 220 ⨯ 220 mm; slice thickness, 3.0 mm; gap, 1.0 mm; b-value, 1000 s/mm^2^. Protocols of optional axial T1-weighted image and axial T2-weighted image sequences are summarized in Table S2 in the <u>Data Supplement</u>.

### Statistical analyses

To assess the performance of the triage research application, sensitivity, specificity, and the area under the receiver operating characteristic curve (AUROC) were calculated. To overcome potential bias caused by an imbalanced dataset, additional statistical values including the area under precision (e.g. sensitivity)-recall (e.g. positive predictive value) curve (AUPRC), and maximum F1 score were analyzed ^11^. For the inter-reviewer agreements of the three radiologists, the Fleiss Kappa was defined as follows; poor agreement, < 0.2; fair agreement, 0.2 ≤ and < 0.4; moderate agreement, 0.4 ≤ and < 0.6; good agreement, 0.6 ≤ and < 0.8; and excellent agreement,≥ 0.8. Using the Student’s t-test, the baseline characteristics were compared between positive and negative AIS groups according to mandatory sequence alone and with the addition of optional sequences. All statistical analyses were performed using MedCalc (version 20.215, MedCalc Software Ltd, USA). *p* < 0.05 was considered to indicate a statistically significant difference.

## Results

### Study Sample characteristics

A total of 831 individuals (mean age, 64 years ± 16; 399 men, 432 women) were enrolled in the study. A flow diagram of the inclusion and exclusion process is illustrated in Figure 1 and the characteristics of the final study population are summarized in Table 1. In total, 201 (mean age, 69 ± 14[SD]; 123 men, 78 women) were positive for AIS, while 630 (mean age, 62 ± 16[SD]; 276 men, 354 women) were negative. The group with AIS was significantly older than the control group and AIS also occurred significantly more frequently in men than women (*p* < 0.001). However, there was no predilection of age or sex between the mandatory sequence alone group and optional sequences groups (p > 0.05).

**Table 1:**
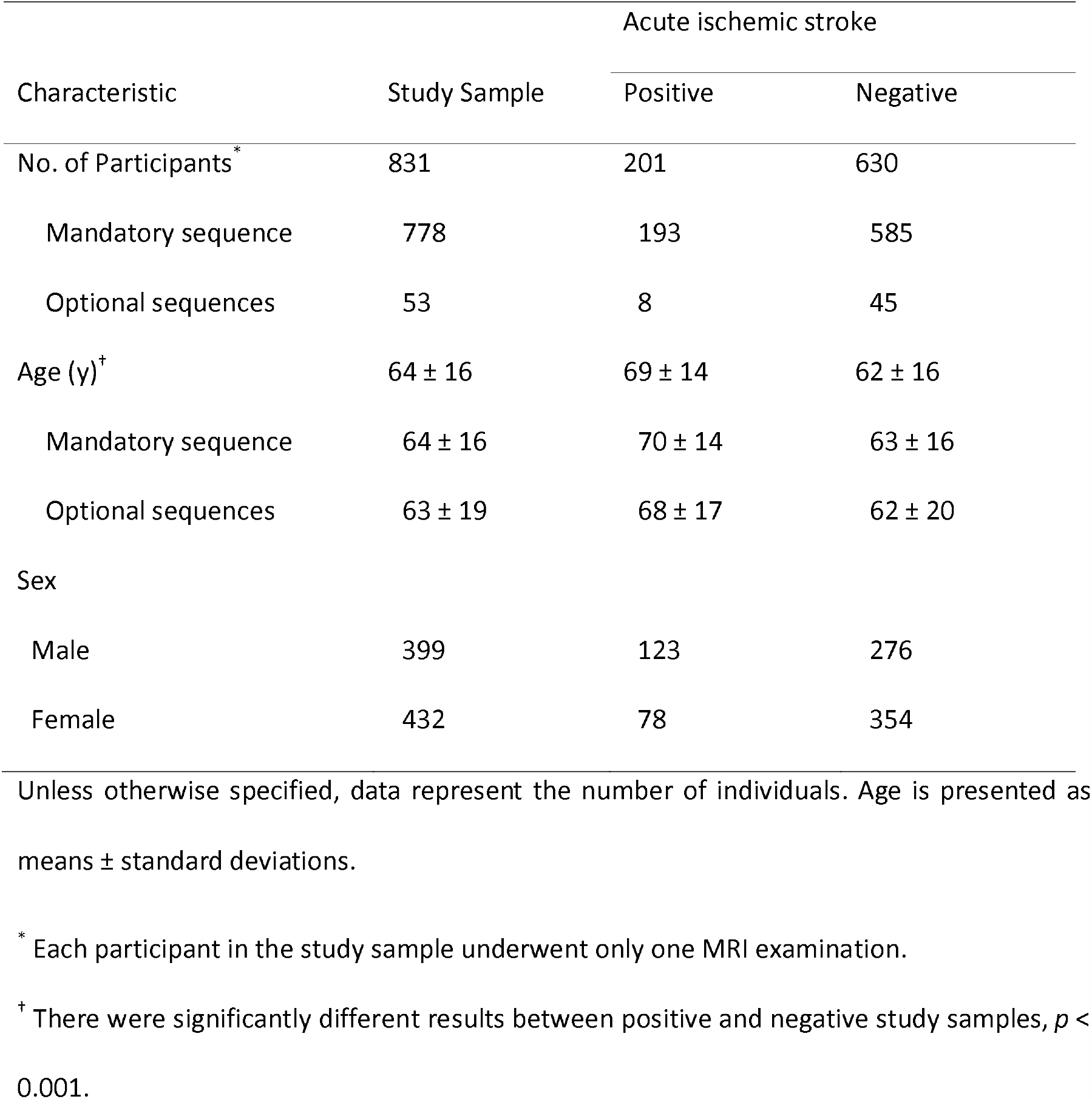
Baseline characteristics of the study sample.

**Figure 1:**
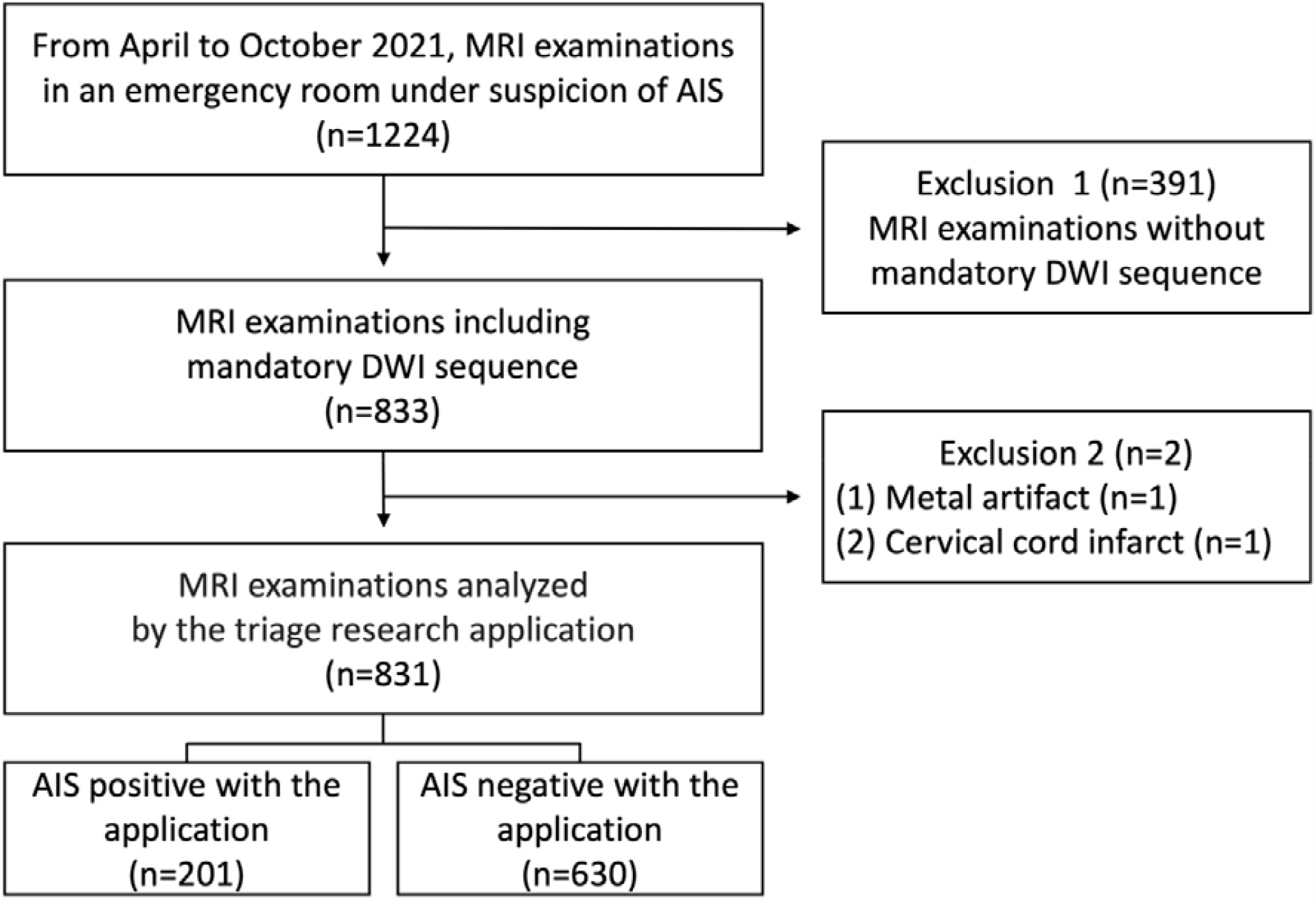
Flowchart of the study sample enrollment. MRI, magnetic resonance imaging; AIS, acute ischemic infarct; DWI, diffusion-weighted image.

### Inter-reviewer agreement

The inter-reviewer agreements among the neuroradiologists for detecting AIS were calculated. The Kappa value for AIS was 0.94 (95% CI, 0.90-0.98, *p* < 0.001), indicating excellent agreement. The Kappa value for the mandatory sequence alone and optional sequences group was 0.85 (95% CI, 0.69-1.00, *p* < 0.001) and 0.94 (95% CI, 0.90-0.98, *p* < 0.001), respectively, indicating excellent agreement. The Kappa value from the sub-analysis for lacunar AIS and non-lacunar AIS was 0.89 (95% CI, 0.85-0.94, *p* < 0.001) and 0.94 (95% CI, 0.90-0.98, *p* < 0.001), respectively, indicating excellent agreement.

### Performance for detecting AIS

The statistical values of the triage research application were as follows; sensitivity, 90%; specificity, 89%; AUROC, 0.95 (95% CI, 0.93-0.96); AUPRC, 0.91 (95% CI, 0.86-0.94); and maximum F1 score, 0.87. The average reporting time from uploading images to presenting results was 61 ± 31 seconds. In terms of the contribution of optional sequences, all statistical values were higher with the mandatory sequence alone than with the addition of optional sequences. Table 2 summarizes the results of this application, and Figure 2 shows the AUROC and AUPRC. Also, illustrations of cases in which the application identified AIS as true positives are presented in Figure 3.

**Table 2:** Detection performance of the triage research application for AIS, with the mandatory sequence alone and the addition of optional sequences.

| Statistical Values | Overall | Mandatory sequence | Optional sequences |
| --- | --- | --- | --- |
| Sensitivity (%) | 89.55 | 89.64 | 87.50 |
| Specificity (%) | 89.05 | 89.40 | 84.44 |
| AUROC (95% CI) | 0.95 (0.93-0.96) | 0.95 (0.93-0.97) | 0.88 (0.76-0.95) |
| AUPRC (95% CI) | 0.91 (0.86-0.94) | 0.91 (0.86-0.94) | 0.88 (0.46-0.98) |
| Maximum F1 score | 0.87 | 0.87 | 0.82 |

This table shows the results of the triage research application for acute ischemic stroke overall, with the mandatory sequence alone, and with the addition of optional sequences; AUROC, area under receiver operating characteristic curve; AUPRC, area under the precision-recall curve; CI, confidence interval.

**Figure 2:**
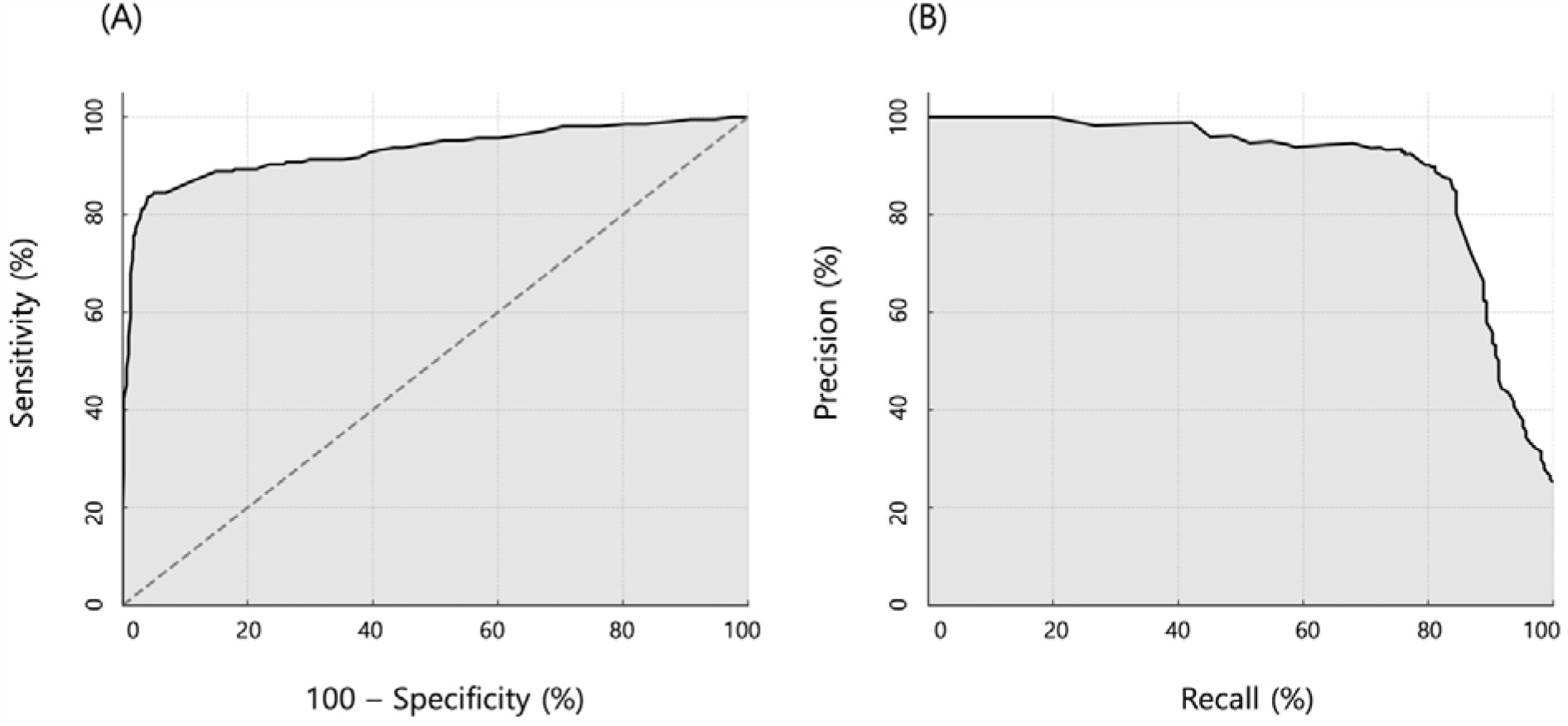
**A.** Receiver operating characteristic curve of the triage research application. **B**. Precision-recall curve of the triage research application.

**Figure 3:**
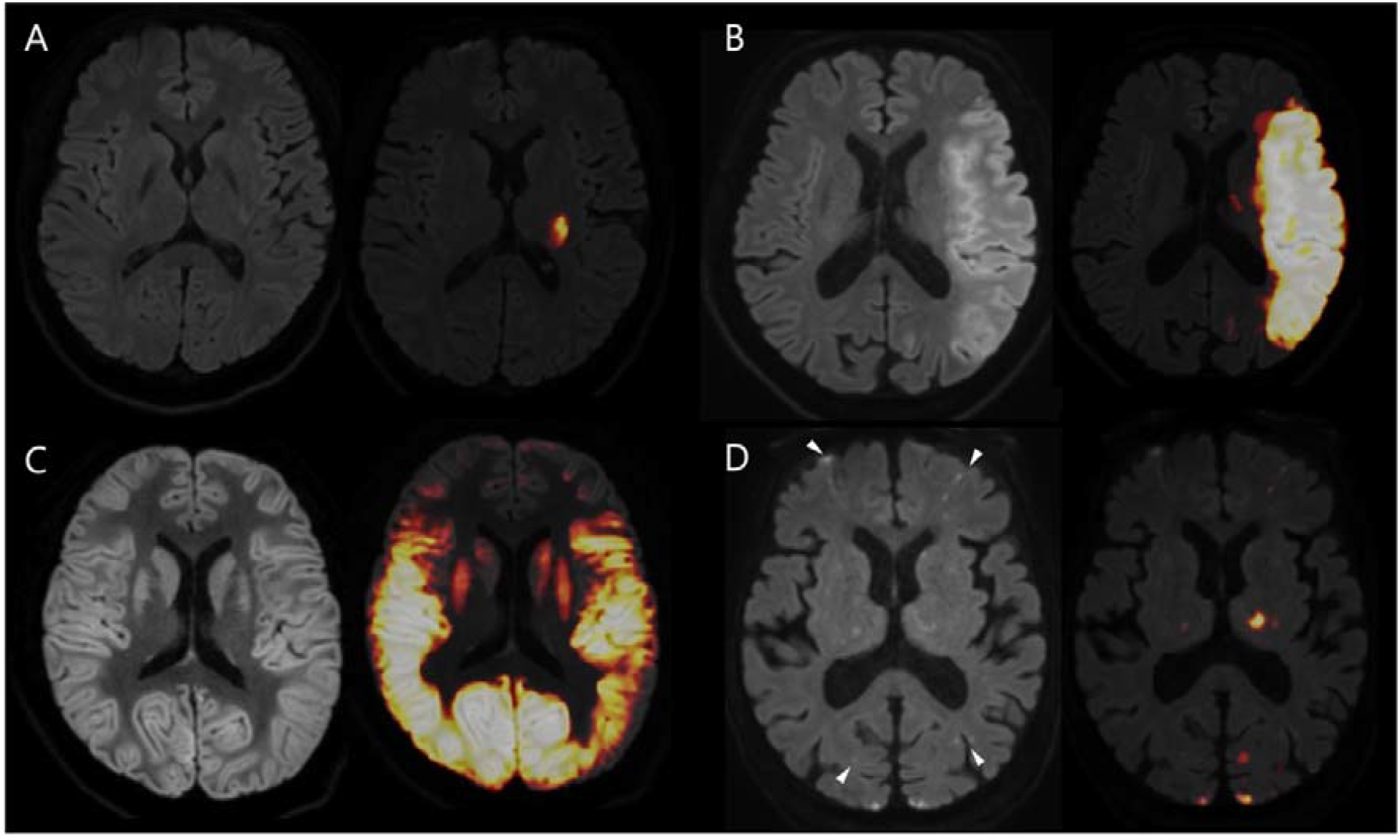
Examples of AIS detected by the triage research application. From left to right, axial DWI, and heat map images are displayed. **A.** Lacunar AIS in the left basal ganglia. **B.** Non-lacunar AIS in the left middle cerebral artery. **C.** Hypoxic ischemic encephalopathy with diffusely elevated DWI signal along the bilateral cerebral cortex and deep gray matter with cerebral parenchymal edema. On the heat map, the application partially identified these features. **D.** Embolic AIS in both hemispheres. On the heat map, the application partially identified these features. The white arrowheads in DWI indicate missing lesions.

### Sub-analysis of performance for detecting lacunar and non-lacunar AIS

The proportion of lacunar AIS and non-lacunar AIS was 31% (63 of 201) and 69% (138 of 201), respectively. This sub-analysis demonstrated a lower sensitivity of 83% for detecting lacunar AIS compared to a sensitivity of 93% for non-lacunar AIS. The sub-analysis showed higher performance for non-lacunar AIS than lacunar AIS. More detailed results are summarized in the <u>Data Supplement</u> with Table S3.

## Discussion

In this study, the performance of the triage research application to detect AIS in an actual emergency department was assessed. The performance of this application in real-world emergency care showed equal or lower sensitivity, specificity, and AUROC compared to a previous validation study ^7^. These differences might be due to the disparate incidence rates of AIS in our institutional emergency room and the previous study rather than from the distinct performance of this application ^7^. Therefore, we additionally calculated the AUPRC and maximum F1 score to mitigate the potential misinterpretation of our results due to an unbalanced study sample ^7,12,13^. The AUPRC and maximum F1 score indicate that the triage research application shows relatively excellent performance for detecting AIS.

When comparing the performance of the mandatory sequence alone and adding optional sequences, analyses based on additional optional sequences presented lower statistical values than those based on the mandatory sequence alone. This result could also be biased due to a possible unbalanced study sample between sequences. However, considering that the optional sequences group showed lower values for both the AUPRC and maximum F1 score, the contribution of adding optional sequences may not improve performance at all, and reduce the performance of the application for detecting AIS instead. Sub-analyses of the application showed that all statistical values for detecting non-lacunar AIS were higher than for lacunar AIS. These results are consistent with a previous validation study ^7^. This is probably due to the difficulties that arose when the triage research application tried to identify relatively small lesions because this application was developed based on the contrast of images from MRI ^7^. Thus, the low detection performance for lacunar AIS also needs to be resolved, despite lacunar AIS not having a significant effect on patient outcomes and being difficult for even experienced radiologists to detect ^14-16^.

The present study has several limitations. Due to the retrospective design of the study, the characteristics of the patient sample may have been biased. However, this limitation was inevitable due to the nature of the study design. Second, because we included MRI examinations from the same vendor at a single institution, we could not generalize the performance of this application across institutions or vendors. Also, all MRI examinations were performed with 3T MRI machines. Research that encompasses a variety of circumstances including different tesla machines is required to overcome this bias. Third, there were no values that could be compared when interpreting the AUPRC and maximum F1 score. Therefore, we could not narrow down the potential cause for different results to different incidence rates of AIS or existing differences in the performance of the application. Nevertheless, these results of our clinical study are thought to provide a milestone for future validation and development of this application. Finally, this study was limited to assessing the application’s detection performance and did not evaluate its far-reaching effects on patient outcomes. While the study results provide valuable insight into the performance of the triage research application, further research is also required to evaluate how its use will impact patient outcomes and to what degree. Future research must incorporate larger and more diverse patient populations under various environmental conditions before our results can be generalized.

## Conclusions

This study has demonstrated that this triage research application, NeuroTriage, can accurately identify AIS on brain MRI, even though its performance was equal or lower to a previous validation study. The addition of optional sequences was not necessary to enhance its detection performance. If this application is utilized, it could lead to the standardization of a faster and more consistent stroke protocol for the emergency room. Therefore, radiologists and clinicians must have a comprehensive understanding of the capabilities and limitations of this triage research application for optimal patient diagnosis.

## Data Availability

More information about its development and validation, and the open-source code of the initial version of the triage research application, NeuroTriage, are available at DOI: https://doi.org/10.1038/s41598-021-86022-7. The source code of the current version of this application cannot be disclosed due to the copyright policy of Siemens Healthineers.

## Abbreviations

AIS: Acute ischemic stroke
DWI: diffusion-weighted image
ADC: apparent diffusion coefficient
AUROC: area under the receiver operating characteristic curve
AUPRC: area under the precision-recall curve.

## Acknowledgements

This study was supported by the research support center of Eunpyeong St. Mary’s Hospital. We are grateful to Siemens Healthineers for providing a work-in-progress application for our use. More information about its development and validation, and the open-source code of the initial version of the triage research application, NeuroTriage, are available at DOI: https://doi.org/10.1038/s41598-021-86022-7. The source code of the current version of this application cannot be disclosed due to the copyright policy of Siemens Healthineers.

