## Supplemental material 1, 2 and 3 for "Assessment of Deep Learning-Based Triage Application for Acute Ischemic Stroke on Brain MRI in the Emergency Room"

### **Supplemental Material 1** *Sample size calculation*

The sample size for a binary classifier was determined using the standard formula used to calculate the sample size for proportions. We determined the sample size using a level of statistical significance of 0.05 and a level of statistical power of 0.80. Based on a previous study [7], the sensitivity was set to 92%, specificity to 88%, and incidence of the population to 4.4%. The calculated sample size for this study was 352.8 individuals. More patients were enrolled in this study than the calculated sample size.

### **Supplemental Material 2** *Inter-reviewer agreement*

The Kappa value for lacunar AIS and non-lacunar AIS was 0.89 (95% CI, 0.84-0.93, $p < 0.001$) and 0.99 (95% CI, 0.90-0.99, $p < 0.001$), respectively, indicating excellent agreement.

### **Supplemental Material 3** *Detection performance for lacunar and non-lacunar AIS.*

The number of lacunar and non-lacunar AIS was 63 and 138, respectively. The sensitivity, specificity, AUROC, AUPRC, and maximum F1 score of the triage research application was 83%, 89%, 0.91, 0.77 and 0.79 for detecting lacunar AIS, and 93%, 89%, 0.96, 0.91, and 0.87 for detecting non-lacunar AIS, respectively. All statistical values were higher for detecting non-lacunar AIS than lacunar AIS.
