## Supplemental Table S1, S2 and S3 for "Assessment of Deep Learning-Based Triage Application for Acute Ischemic Stroke on Brain MRI in the Emergency Room"

**Table S1:** Detection performance for AIS in a previous validation study.

| Statistical Measure | Internal Validation Test | External Validation Test |
|---|---|---|
| No. of participants | 1050 | 1072 |
| Sensitivity (%) | 92 | 90 |
| Specificity (%) | 88 | 97 |
| AUROC | 0.95 | 0.97 |

**Table S2:** MRI protocols for optional sequences.

| Protocol parameters | T1-weighted imaging | T2-weighted imaging |
|---|---|---|
| Repetition time (ms) | 2090 | 6310 |
| Echo time (ms) | 3 | 100 |
| No. of excitations | 1 | 2 |
| Matrix size | 256 × 240 | 480 × 336 |
| Flip angle (°) | 9 | 150 |
| Field of view (mm) | 210 × 210 | 210 × 210 |
| Slice thickness (mm) | 3.0 | 4.0 |
| Gap (mm) | 4.0 | 5.0 |

3T MRI scanner (MAGNETOM Vida, Siemens Healthineers, Erlangen, Germany) with a 64-channel. A head & neck coil was applied for optional sequences.

**Table S3:** Detection performance for lacunar and non-lacunar AIS.

| Statistical Measure | Lacunar AIS | Non-lacunar AIS |
|---|---|---|
| No. of participants | 63 | 138 |
| Sensitivity (%) | 82.54 | 92.75 |
| Specificity (%) | 89.05 | 89.05 |
| AUROC (95% CI) | 0.91 (0.89-0.93) | 0.96 (0.95-0.98) |
| AUPRC (95% CI) | 0.77 (0.65-0.86) | 0.91 (0.85-0.95) |
| Maximum F1 score | 0.79 | 0.87 |

AIS, acute ischemic stroke; AUROC, area under receiver operating characteristic curve; AUPRC, area under precision-recall curve; CI, confidence interval.
