## Supplemental Figure S1, S2 and S3 for "Assessment of Deep Learning-Based Triage Application for Acute Ischemic Stroke on Brain MRI in the Emergency Room"

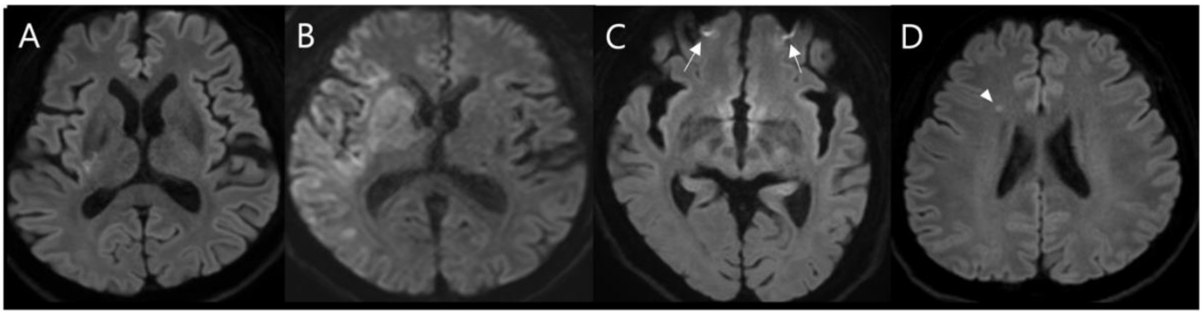


**Figure S1**: Example of axial diffusion-weighted imaging (DWI) for acute ischemic stroke (AIS) and potential artifacts. **A.** Lacunar AIS in the right basal ganglia. **B.** Non-lacunar AIS in the right middle cerebral artery territory. **C.** Susceptibility artifacts in both frontal hemispheres caused by magnetic field distortion (white arrows). **D.** T2 shine-through artifacts in the right corona radiata (white arrowhead).

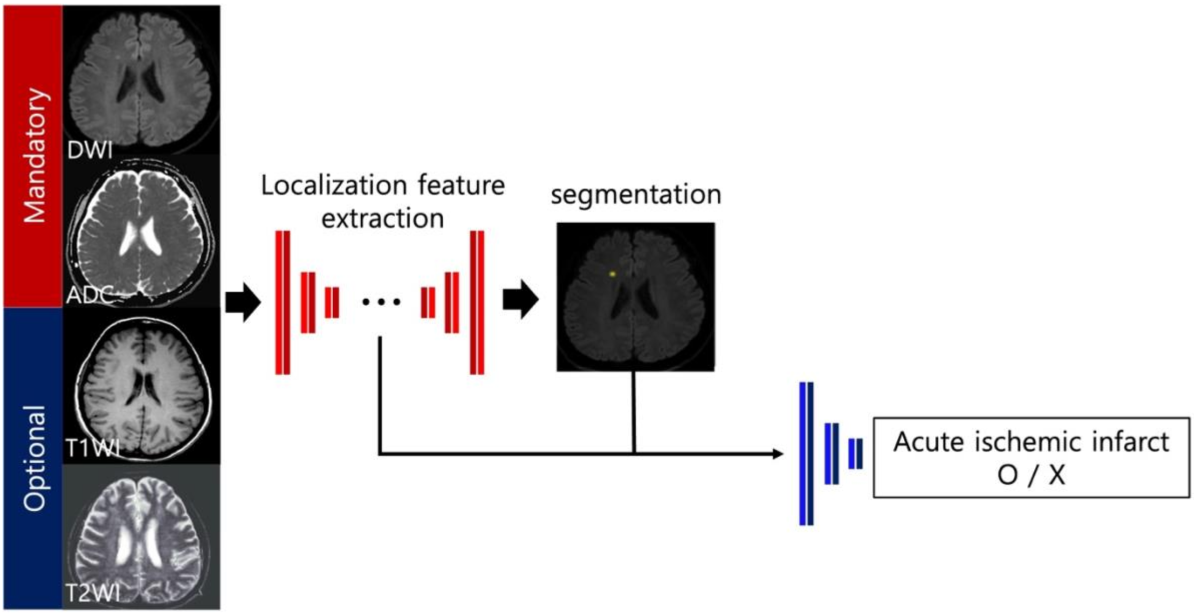


**Figure S2:** The network architecture of the triage research application. 3D convolutional neural network was applied to the AIS detection process based on the mandatory sequence, the DWI with the apparent diffusion coefficient map. As a result of the detection process, the application provides the AIS score in a patient-wise manner and the heat map.

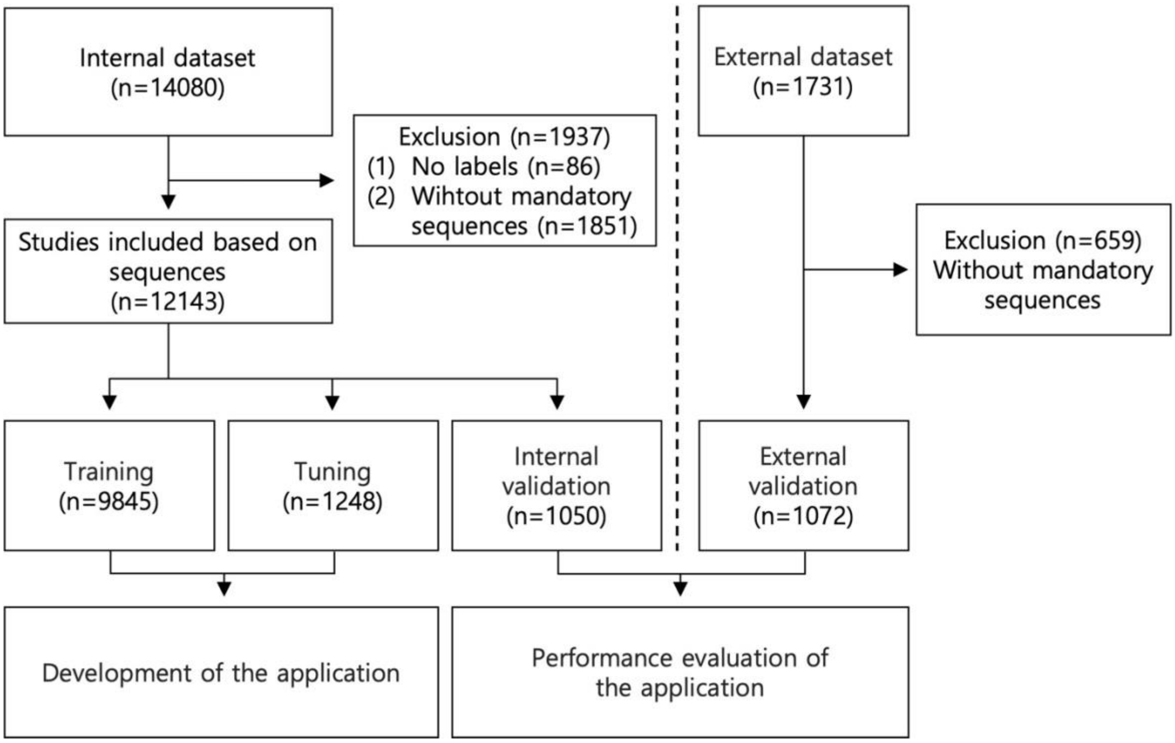


**Figure S3:** Flowchart of the development of the triage research application and its performance evaluation using an internal and external dataset.
